# Lexical meaning is lower-dimensional in psychosis: the intrinsic geometry of the semantic space

**DOI:** 10.1101/2025.06.03.25328940

**Authors:** Claudio Palominos, Frederike Stein, Tilo Kircher, Rosa Ayesa-Arriola, Lena Palaniyappan, Philipp Homan, Iris E. Sommer, Wolfram Hinzen

**Affiliations:** Department of Translation & Language Sciences, Universitat Pompeu Fabra, Barcelona, Spain; Department of Psychiatry and Psychotherapy, University of Marburg, Germany; Center for Mind, Brain and Behavior (CMBB), Marburg, Germany; Department of Psychiatry, Marqués de Valdecilla University Hospital, IDIVAL, School of Medicine, University of Cantabria, Santander, Spain; Biomedical Research Center in Mental Health Network (CIBERSAM), Health Institute Carlos III, Madrid, Spain; Douglas Mental Health University Institute, Department of Psychiatry, McGill University, Montreal, Quebec, Canada; Department of Medical Biophysics, Schulich School of Medicine and Dentistry, Western University, London, Ontario, Canada; Department of Psychiatry, Schulich School of Medicine and Dentistry, Western University, London, Ontario, Canada; Robarts Research Institute, Schulich School of Medicine and Dentistry, Western University, London, Ontario, Canada; Department of Adult Psychiatry and Psychotherapy, University of Zurich, Zurich, Switzerland; Neuroscience Center Zurich, University of Zurich and ETH Zurich, Zurich, Switzerland; Department of Neuroscience, University Medical Center Groningen, Antoni Deusinglaan 2, room 117 Groningen, Netherlands; Institució Catalana de Recerca i Estudis Avançats (ICREA), Barcelona, Spain

**Keywords:** Schizophrenia, Psychosis, Language Models, Semantics, Word embeddings

## Abstract

Diverse language models (LMs), including large language models (LLMs) based on deep neural networks have come to provide an unprecedented opportunity for mapping out the semantic spaces navigated in speech and their distortions in mental disorders. Recent evidence has pointed to higher mean semantic similarities between words in psychosis, conceptualized as a ‘shrunk’ (more compressed) semantic space. We hypothesized that the high dimensionality of the vector spaces defined by the embeddings of speech samples through LMs would also be easier to reduce in psychosis. To test this, we used principal component analysis (PCA) to calculate different metrics serving as proxies for reducibility, including the number of components needed to reach 90% of variance, and the cumulative variance explained by the first two components. For further exploration, intrinsic dimensionality (ID) was also estimated. Results confirmed significantly higher reducibility of the semantic space in psychosis across all measures and three languages. This result points to the existence of an underlying intrinsic geometry of semantic associations during speech, which may underlie more surface-level measurements such as semantic similarity and illustrates a new foundational approach to speech in mental disorders.

## 1. Introduction

When communicating, we draw concepts from our semantic memory into structured speech, connecting different words and phrases to convey meaning. Through this process, we metaphorically navigate a semantic space, traversing different regions in a trajectory that as such gives meaning to the discourse as a whole. This notion aligns with the framework of distributional semantics in language models (LMs), where (sub)words, sentences, or even larger linguistic units are represented as vector embeddings in a high-dimensional space. Metrics such as cosine similarity capture the relationships between vectors: the smaller the angle between them, the more semantically related the words, sentences, or texts are, as they tend to occur in similar contexts in the training data used to build these models.

Several semantic similarity metrics derived from embeddings have been employed to analyze spontaneous speech of people with schizophrenia spectrum disorder (SSD), who exhibit distinct navigational patterns through semantic space compared to healthy controls (Alonso-Sánchez et al., 2022; Ciampelli et al., 2023; He et al., 2024a; Palominos et al., 2024; Nour et al., 2023). These differences have informed the development of classification models aimed at distinguishing clinical groups (Bedi et al., 2015; Voppel et al., 2021). At the core of this research program lies the question of whether major psychiatric disorders such SSD disrupt the underlying organization of the semantic space and the way it manifests in natural speech (Kircher et al., 2018).

Yet despite the growing use of semantic variables in recent studies, little is known about how these variables relate to each other, or whether they reflect overlapping or independent aspects of semantic organization. This lack of integration complicates efforts to interpret findings across studies or to identify robust, generalizable features of semantic disruption in psychosis. A preliminary attempt to address this was made in Palominos et al. (2025), where a composite index was constructed by aggregating over a hundred semantic features derived from language models. However, a deeper understanding of the structure and dimensionality of these embeddings remains lacking.

This study aims to capture an aspect of the geometry of the high-dimensional space of embeddings derived from LMs and spontaneous speech in SSD. Specifically, we infer the geometry of a given speech sample using principal component analysis (PCA) and an estimation of intrinsic dimensionality (ID). While PCA finds an orthogonal basis in a given coordinate system that best explains variance in the data, ID identifies the minimum number of degrees of freedom needed to describe the data without significant loss of information (Fukunaga, 2013). The number of principal components (PCs) required to explain a given amount of variance, along with the variance explained by the first components, and the values if ID, serve as a measure of the reducibility of the embeddings and how semantically associated the words are. When comparing two speech samples of equal length, a smaller number of components or a lower ID suggests that the shared information across the set of embeddings within each speech is more redundant, as it can be captured using fewer dimensions.

While the focus is our analysis is primary lexical, ultimately lexical selection is entangled in speech with grammatical structure. Content words do not occur in isolation but are embedded in syntax, meaning that these measures may also reveal aspects of grammatical organization. This interdependence justifies the exploration of ID across different layers when using BERT, a contextual large language model, where early layers capture more surface features, middle layers syntactic information and later ones encode more abstract and semantic relationships (Jawahar et al., 2019). Examining how ID varies across BERT layers may help uncover how grammatical and semantic constraints shape the latent structure of language in SSD.

We hypothesized that the intrinsic geometry of the semantic space in SSD is lower-dimensional compared to that of HC, which aligns and complements the observed patterns of a ‘shrunk’ of semantic space as manifest through increased mean semantic similarity (He et al., 2024a; Çabuk et al., 2024; Arslan et al., 2024), or a reduced convex hull volume in SSD (Palominos et al., 2024). In line with this hypothesis, we found across three datasets and three languages that SSD speech is more reducible compared to healthy controls, whether using PCA or ID. This result illustrates a new principled approach, focussed on the geometry of the semantic space through observable word embeddings, which may help to explain the previously observed pattern of a ‘shrunk’ (more compressed) semantic space, which becomes more semantically interconnected in SSD.

## 2. Methods

### 2.1 Data collection

The datasets comprised speech samples from German (N=86), Spanish (N=84) and English (N=94) speakers. Demographics and clinical data are summarized in Table 1, including the different compositions of subsamples. In the English sample, SSD was divided into first-episode psychosis (FEP) and chronic schizophrenia (CS) to examine potential differences within SSD, with clinical high-risk (CHR) group also included. In each language, the samples were obtained through the same task of describing pictures from the Thematic Apperception Task (TAT). This test provided the opportunity to control that participants describe specific topics, limiting thematic variability. More details of related German, Spanish, and English samples can be found in Palominos et al., 2024, He et al., 2024b, and He et al., 2024a, respectively. For all the samples, data collection, protocol and methodology were approved by the local ethical committees (German: Ethik-Kommission des Fachbereichs Humanmedizin der Philipps-Universität Marburg (AZ 07:14); Spanish: CEIm internal code 2021.119; English: The Research Ethics Committee of University of Western Ontario, London, Canada (Project ID: 108268; Most recent review reference: 2022-108268-71496; Study Title: The Pathophysiology of Thought Disorder in Psychosis (TOPSY)).

**Table 1:** Demographics of participants and comparisons for each sample.

| German | HC | SSD | HC – SSD |
| --- | --- | --- | --- |

| | (mean ± SD) | (mean ± SD) | (p-value) |
| --- | --- | --- | --- |
| N° participants | 44 | 42 |  |
| Age (years) | 42.9 ± 12.3 | 40.5 ± 11.9 | 0.3766 |
| Female (%) | 32% | 38% | 0.7008 |
| Education (years) | 14.7 ± 3.3 | 11.9 ± 2.0 | 0.0000*** |
| IQ | 117.5 ± 14.2 | 111.6 ± 15.9 | 0.0786 |
| Age of onset (years) | - | 17.7 ± 8.2 | - |

| | HC<br>(mean ± SD) | SSD<br>(mean ± SD) | HC - SSD<br>(p-value) |
| --- | --- | --- | --- |
| **Spanish** |  |  |  |
| N° participants | 41 | 43 |  |
| Age (years) | 41.3 ± 7.8 | 38.1 ± 9.0 | 0.0844 |
| Female (%) | 41.5% | 48.8% | 0.6459 |
| Education (years) | 11.6 ± 2.6 | 10.5 ± 2.0 | 0.1145 |

| | HC<br>(mean ± SD) | CS<br>(mean ± SD) | CHR<br>(mean ± SD) | FEP<br>(mean ± SD) |
| --- | --- | --- | --- | --- |
| **English** |  |  |  |  |
| N° participants | 29 | 18 | 18 | 29 |
| Age (years) | 21.5 ± 3.6 | 28.5 ± 7.4 | 22.4 ± 4.1 | 21.4 ± 2.2 |
| Female (%) | 24.1% | 22.2% | 27.7% | 27.6% |

|  | HC – CS<br>(p-value) | HC – CHR<br>(p-value) | HC – FEP<br>(p-value) |
| --- | --- | --- | --- |
| Age (years) | 0.0000*** | 0.4464 | 0.8606 |

|  | CS – CHR<br>(p-value) | CS – FEP<br>(p-value) | CHR – FEP<br>(p-value) |
| --- | --- | --- | --- |
| Age (years) | 0.0047 | 0.0000*** | 0.2817 |

*Note:* IQ scores are derived from the MWT-B scale, which assesses verbal premorbid intelligence. Gender comparisons were performed using the $\chi^2$ test, while t-tests were used for comparing other variables. Significance levels are denoted as follows: \* $p < .05$ , \*\* $p < .01$ , \*\*\* $p < .0001$ .

### 2.2 Word embeddings

300-dimensional word embeddings for each speech sample were obtained using fastText (Grave et al., 2018), and 768-dimensional embeddings were obtained using BERT (Bidirectional Encoder Representations from Transformers; Devlin et al., 2019). When fastText was used, unique content words were retained to avoid measuring effects related to lexical diversity, while stopwords and punctuation were removed. However, in a post-hoc analysis, all content words were retained. Preprocessing was carried out using spaCy (version 3.6.0), and the models ‘de_core_news_lg’ for German, ‘es_core_news_lg’ for Spanish, and ‘en_core_web_lg’ for English were applied. When BERT was used, all words were retained, and the models ‘dbmdz/bert-base-german-cased’ for German, ‘dccuchile/bert-base-spanish-wwm-cased’ for Spanish, and ‘bert-base-cased’ for English were employed.

### 2.3 Principal Components Analysis

A principal component analysis (PCA) was conducted on the set of embeddings for each speech sample, with columns as the number of dimensions and rows the number of words in the sample. This allowed each speech sample to be represented by a matrix with fewer dimensions, using the principal components of the word embeddings, instead of the original 300 (or 768 for BERT), while retaining most of the variance. From the PCA, we derived variables indicating the amount of variance explained by the principal components. To do this, we defined *Ncomp*_*X*_ as the number of components required to explain *X* percent of variance, and *EXVar*_*n*_ the cumulative explained variance by the first *n* principal components. For group comparisons, we specifically used *Ncomp*_90_ and *EXVar*_2_, capturing both ends of the components. However, both functions (*Ncomp*_*x*_and *EXVar*_*n*_) proved effective in signalling group differences across different levels of *x*.

### 2.4 Intrinsic Dimensionality

Intrinsic dimensionality (ID) is the lowest number of dimensions required to represent a given dataset without significant information loss. It captures the underlying structure of high-dimensional data and serves as a proxy for reducibility (Fukunaga, 2013). Since linear techniques such as PCA may not effectively capture nonlinear relationships within the data, a variety of alternative dimensionality reduction methods have been proposed (Van der Maaten et al., 2009).

In this study, we estimated the intrinsic dimensionality of the embeddings for each speech sample. To do that we used a Maximum Likelihood Estimation (MLE) method (in Python, *scikit-dimension*) (Levina & Bickel, 2004), which estimates the effective number of dimensions in the correlation structure. The procedure involved first, obtaining the word embeddings for each speech sample, and second, applying MLE to estimate the ID from this correlation structure. This yielded a single ID value for each speech sample. As a post-hoc analysis we also computed the correlation matrix and cosine similarity across embeddings and estimated ID values (see SM).

Given that the lexical concepts of each speech sample refer to a specific picture description, a significant reduction in the intrinsic dimensionality with respect to the original word embeddings is expected, that is, the embeddings will have more information than what is needed for a given picture, and they share semantic properties that allow to reduce this information into a lower dimensional space. In this context, ID provides an estimate of the number of latent degrees of freedom or abstract features required to encode the linguistic patterns within speech, offering insights into the structure of semantic expression.

### 2.5 Statistical analysis

To determine group differences between reducibility variables, we used different models based on the sample, controlling for number of words, education (except the English sample, where Education was a categorical variable), age, and gender in all cases. For the German sample, we ran a mixed-effects model to estimate population-averaged effects controlling for picture effects, which have proved significant in Palominos et al. (2024). In contrast, in Spanish and English samples, we ran a generalized linear model (GLM) averaging the values across pictures. This was necessary after we found that the assumption of normality was violated in these datasets. Finally, to rule out the effect of differing word counts, we not only controlled for this in the models but also ran a t-test to verify that there were no significant differences between groups within each sample (see supplementary material (SM)).

### 2.6 Correlation between measures

Finally, we calculated the pairwise Pearson correlations between all measures and two reference metrics: the average cosine similarity between consecutive words (hereafter, *semantic similarity*), and the dispersion of word embeddings relative to their centroid (hereafter, *dispersion*). Both metrics were computed using *fastText* embeddings, as these two measures have been widely studied in prior research using various models.

## 3. Results

A preliminary analysis showed that *Ncomp*_90_ was strongly correlated with the number of contents words (Pearson correlation, r = 0.905). This strong correlation is anticipated, given that word embeddings are already low-rank representations. However, we confirmed this expected relationship to ensure robustness in subsequent analyses. An equivalent pattern was found between *EXVar*_2_ and the number of content words (Pearson correlation, r = -0.519) which means that with more words less variance is explained by the first components. Both metrics, including related ones (such as various thresholds of variance explained, or the variance accounted for by a certain number of components), indicate an expected decrease in reducibility as the number of content words increases. Although no significant differences were found in the number of words when comparing groups in each sample (Figure S1), the correlation between word count and our metrics supports its inclusion as a covariate in the regression models.

### 3.1 Regression models in different samples

#### Non-contextual model: fastText

Figure 1 presents the model results for fastText model, showing that when controlling by picture, SSD and FEP samples required smaller *Ncomp*_90_. Figure 2 presents equivalent results for the regression of *EXVar*_2_, which shows that the first two components explain more variance in SSD and FEP compared to HC. Taken together, these results suggest that the word embeddings derived from speech in the SSD group are more reducible than those in the HC group. Fewer principal components were required to represent the original 300-dimensional embeddings in SSD compared to HC. An equivalent result is presented in Figure S2 and Figure S3, where all words are preserved rather than only the unique words.

**Figure 1:**
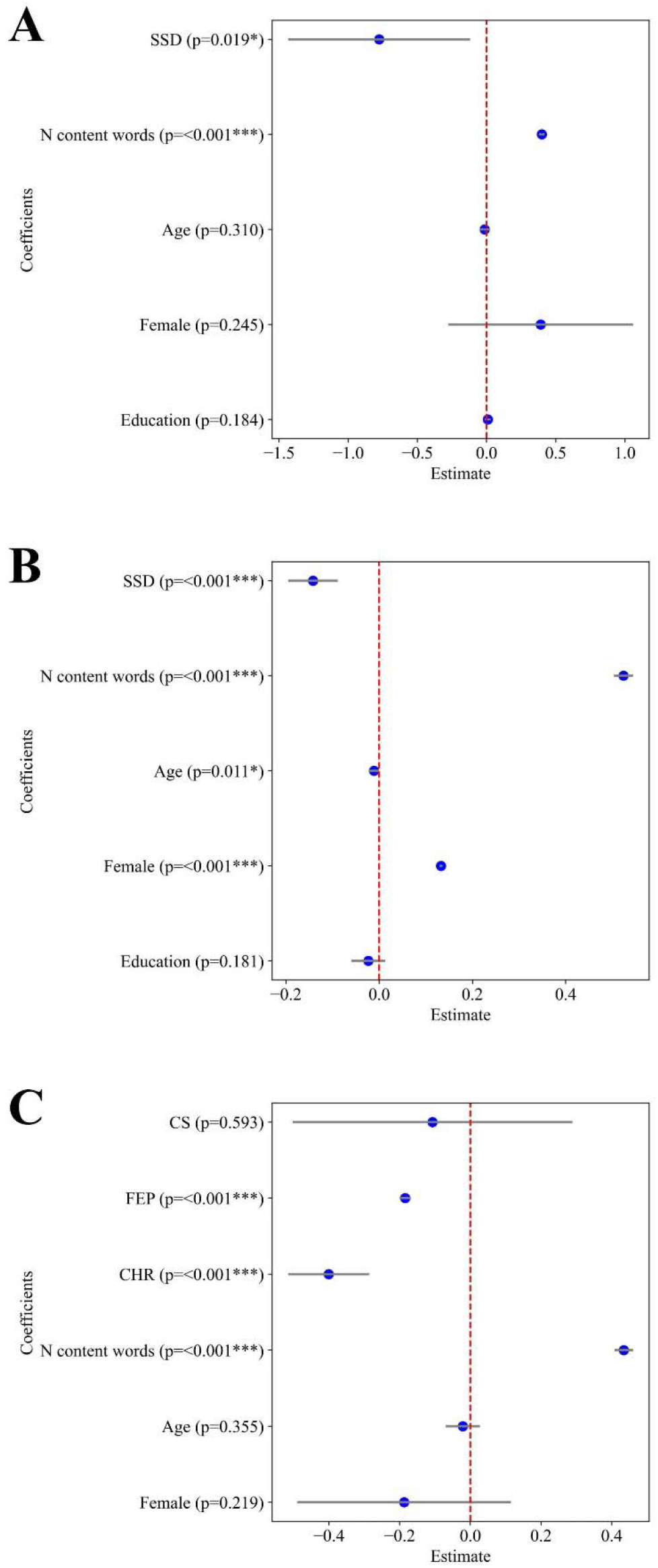
Estimated regressions coefficients for *Ncomp*_90_ across languages. Panel A: German sample; Panel B: Spanish sample; Panel C: English sample. Each blue dot indicates the estimated coefficient, with horizontal lines representing the 95% confidence interval (0.025–0.975). P-values are shown next to each regressor.

**Figure 2:**
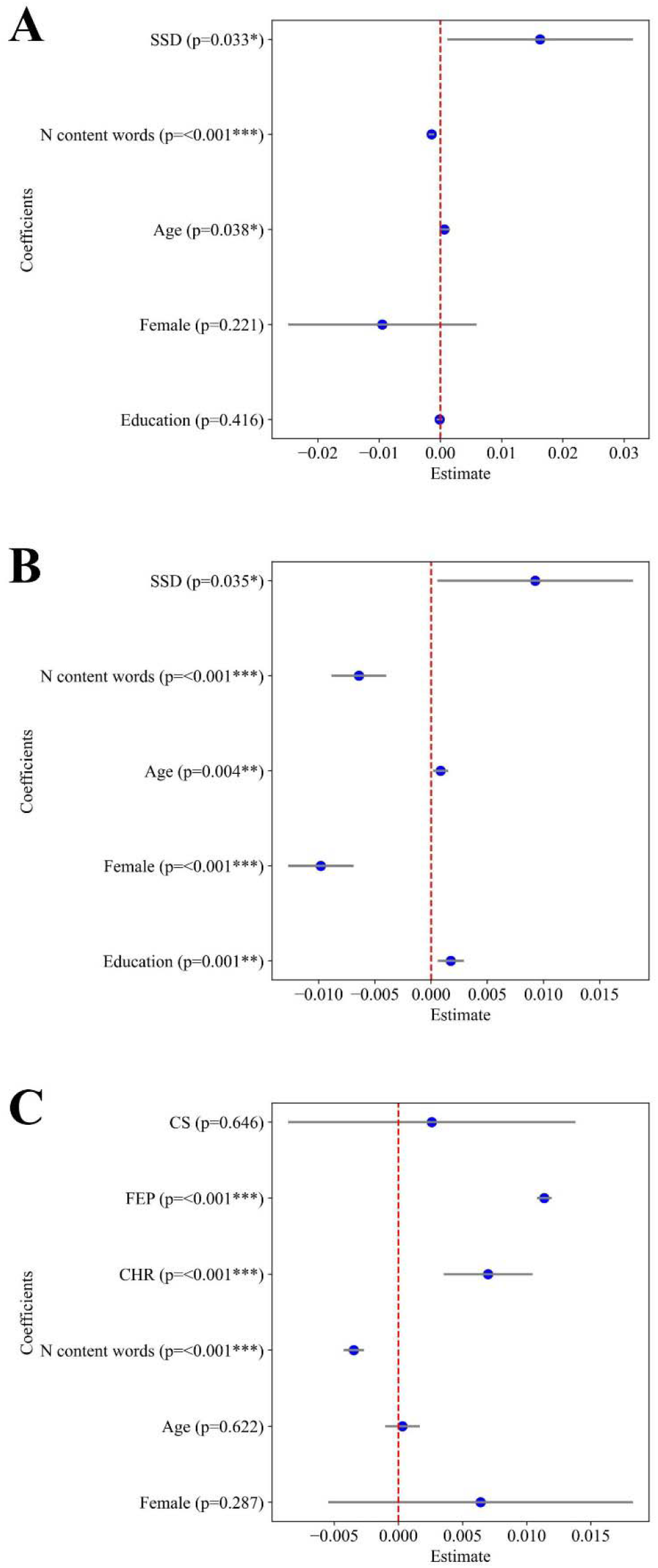
Estimated regressions coefficients for *EXVar*_2_ across languages. Panel A: German sample; Panel B: Spanish sample; Panel C: English sample. Each blue dot indicates the estimated coefficient, with horizontal lines representing the 95% confidence interval (0.025–0.975). P-values are shown next to each regressor.

#### Contextual model: BERT

Figure 3 shows the model results for ID (Panels A, C, E) and *Ncomp*_90_ (Panels B, D, F), both using BERT embeddings. Consistent with previous findings, ID was significantly lower in individuals with SSD compared to HC in both the German and Spanish samples. In the English sample, ID was significantly lower in individuals with FEP compared to HC. However, in the same sample, ID was significantly higher in individuals with CS and CHR compared to HC. This discrepancy suggests a divergence between results obtained using PCA-based measures and those based on ID in the English sample.

**Figure 3:**
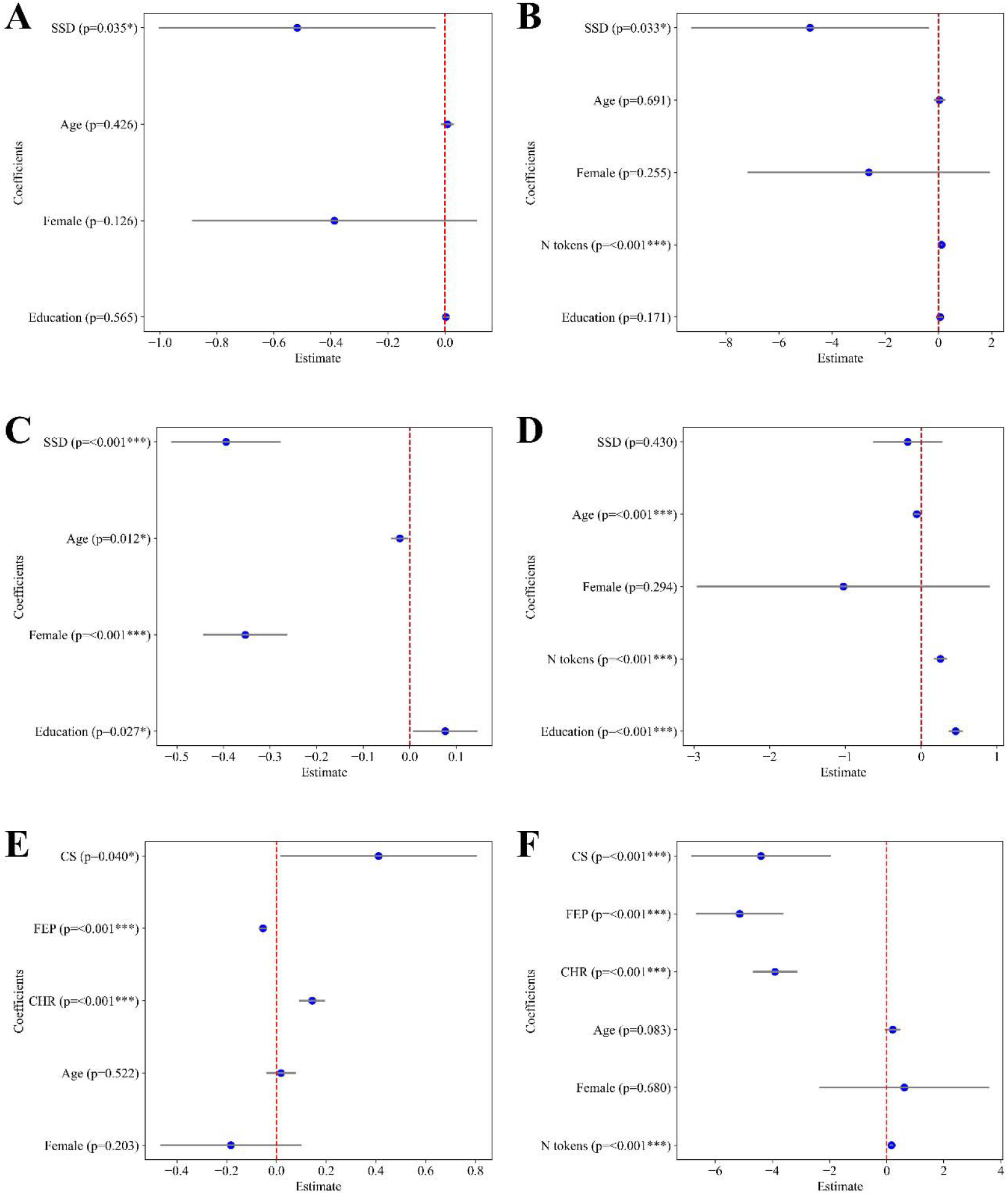
Estimated regression coefficients for ID and *Ncomp*_90_ across languages using BERT embeddings. Panels A and B: German sample; Panels C and D: Spanish sample; Panels E and F: English sample. Left panels (A, C, and E) show estimated coefficients from models regressing ID, while right panels (B, D, F) show the estimated coefficients from models regressing *Ncomp*_90_. Each blue dot indicates the estimated coefficient, with horizontal lines representing the 95% confidence interval (0.025–0.975). P-values are shown next to each regressor.

To further address these differences, *Ncomp*_90_ was found to be significantly lower in all clinical groups compared to HC, except in the Spanish sample, where no significant differences were observed.

### 3.2 Intrinsic dimensionality across BERT layers

Figure 4 shows the average ID estimates for each group across the three languages using BERT-base (12 layers). Notably, only in the German sample significant groups differences appear after layer 6. Overall, ID is significantly lower in SSD groups compared to HC in both German and Spanish samples, and significantly lower in FEP and CHR groups, compared to HC in the English sample. These differences are summarized in Table S1. Equivalent plots for the average ID estimates using correlation and cosine similarity matrices are provided in Figure S4.

**Figure 4:**
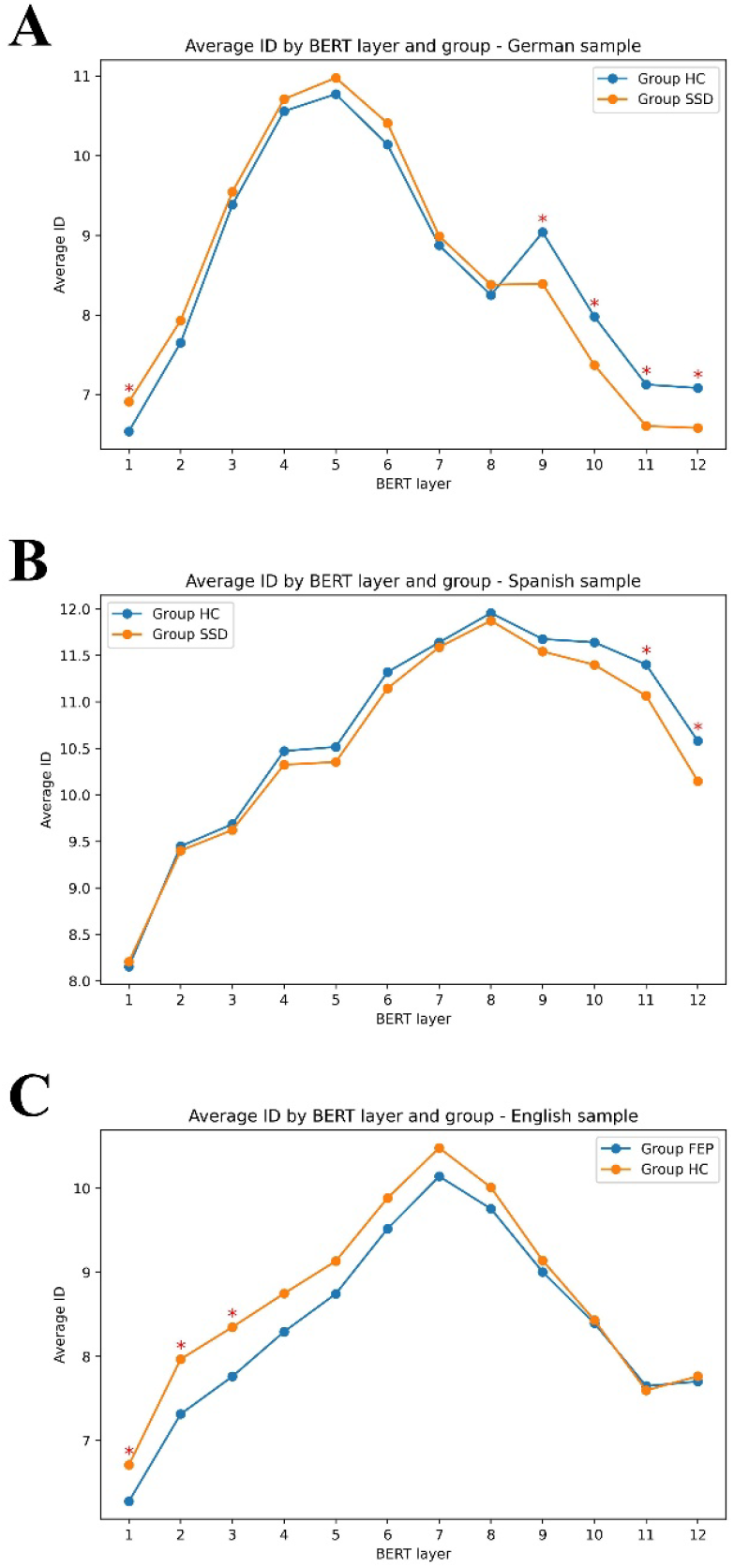
Average Intrinsic Dimensionality (ID) estimates across BERT layers for each group and sample. ID was computed layer-by-layer using 12-layer BERT embeddings for each group. Panel A: German sample; Panel B: Spanish sample; Panel C: English sample. Red asterisks indicate t-test statistically significant group differences at specific layers. Note that in the Spanish sample, only FEP and HC groups are compared here. A plot including all four groups is provided in Figure S5.

### 3.3 Correlation between measures

We calculated Pearson correlations coefficients between five variables: mean semantic similarity, dispersion to the centroid, *Ncomp*_90_, *EXVar*_2_, and *ID*. The resulting correlation patterns for each group are shown in Figure 5. As expected, *Ncomp*_90_ and *EXVar*_2_ are strongly negatively correlated: if fewer components are needed to explain 90% of variance, it is likely that the variance explained by the first two components is higher. This correlation is weaker in the German sample, suggesting that the two measures may be capturing different aspects of reducibility in different samples.

**Figure 5:**
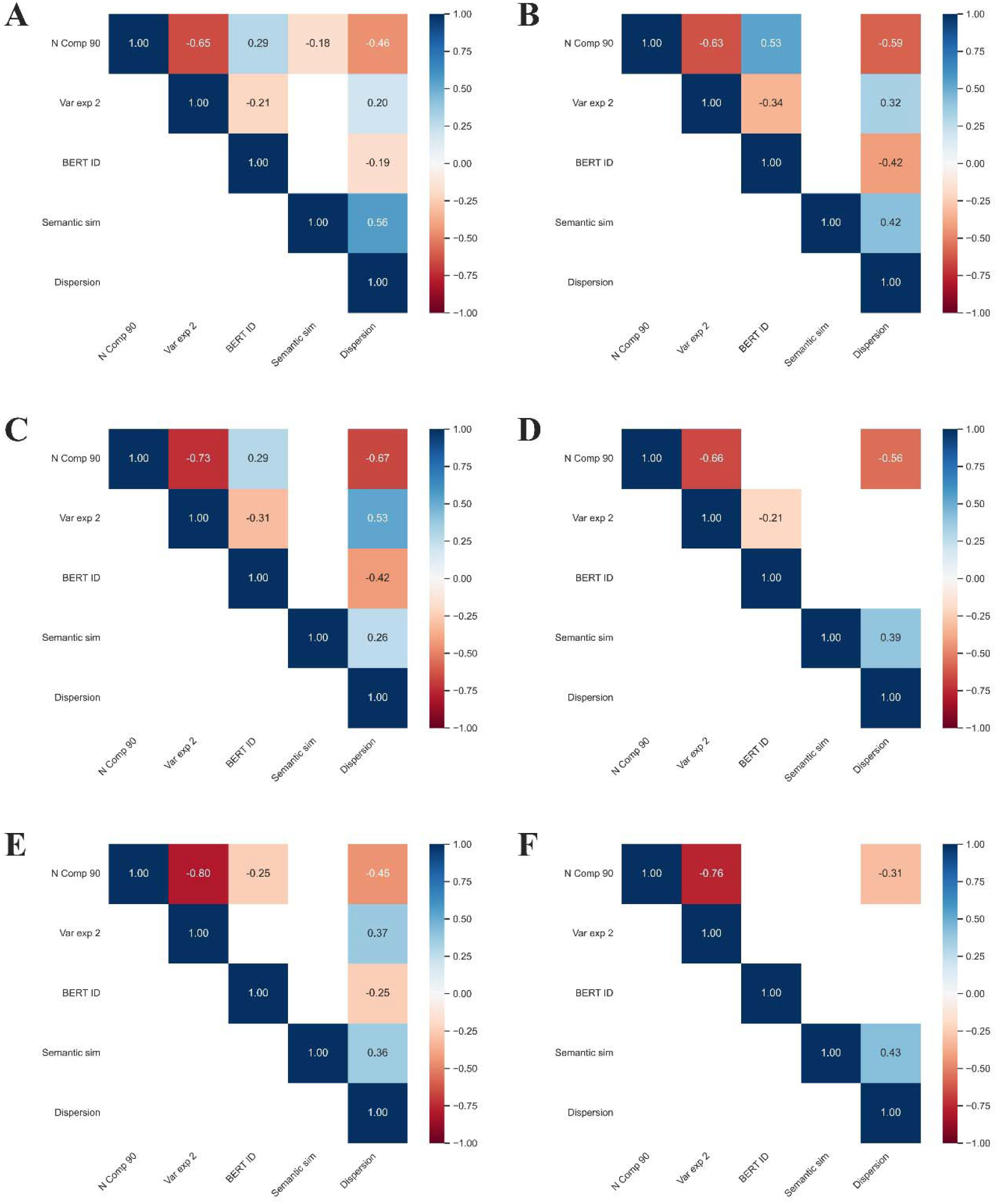
Correlation matrices of variables by language and group.

*Ncomp*_90_ and *ID* were only weakly correlated in the English and Spanish samples, and even weakly negatively correlated in English clinical group. This suggests that *ID*-based reducibility in BERT may capture a different aspect of structure, likely because *ID* includes all tokens in the speech, and therefore is not purely lexical. Similarly, correlations between *EXVar*_2_ and *ID* were weakly negatively correlated, but only in the German and Spanish sample.

In addition, only in the SSD group of the German sample did *Ncomp*_90_ show a weak negative correlation with mean semantic similarity. Likewise, *ID* showed no correlation with mean semantic similarity. While this may indicate a lack of direct relation, it may also point to the need for a more nuance understanding of how both variables interact, specially given that they can be differently affected by speech length.

Finaly, the weak to moderate negative correlation between *Ncomp*_90_ and dispersion contributes to clarify the nature of these relationships. In the context of a picture description task, this may reflect that when more components are needed, indicating richer semantic diversity, the embeddings of lexical words remain closer to the topic. This suggests that they collectively contribute to characterizing the picture being described.

Each heatmap shows pairwise Pearson correlation between variables for German (A-B), Spanish (C-D), and English (E-F) samples. Left panels (A, C, and E) show results for clinical groups (SSD, or FEP in English sample; correlation matrices for other groups (CS, CHR) are presented in Figure S6), while right panels (B, D, and F) show HC. Only statistically significant correlations are shown (p < 0.05), with red indicating negative and blue indicating positive correlations. N Comp 90: Number of principal components required to explain 90% of the variance. Var exp 2: Variance explained by the first two principal components. BERT ID: Intrinsic dimensionality estimated from BERT embeddings. Semantic sim: Mean cosine similarity between consecutive content words in a speech sample, *fastText*. Dispersion: Dispersion of all word embeddings with respect to its centroid, using *fastText*.

## 4. Discussion

Recent studies have shown a pattern of *increase* in semantic similarity measures in SSD compared to HC (Alonso-Sánchez et al., 2022; He et al., 2024a; Çabuk et al., 2024; Arslan et al., 2024), conceptualized as a ‘shrunk’ or more compressed semantic space in SSD. Using word embeddings from both static (fastText) and contextual (BERT) models across three distinct languages (German, Spanish, and English datasets), we found that SSD speech samples were significantly more reducible than those of HC, as measured by a set of variables including the number of principal components required to explain variance, the variance explained by early components, and intrinsic dimensionality (ID). These converging findings support the interpretation that SSD speech exhibit more semantic redundancy and reduced semantic variability.

Although our primary aim was to investigate reducibility in lexical meaning using a non-contextual model like fastText, we extended this approach to a contextual model (BERT), which captures all words, including functional ones, and encodes richer semantic nuances though high-dimensional representations. While some results generalized across models, the patterns were less consistent in the English sample, where only FEP showed significantly less ID, and CS and CHR showed a significantly higher ID compared to HC. Additionally, in the Spanish sample, *Ncomp*_90_ did not differ significantly HC. One possible explanation is that lexical reducibility using fastText may underlie the reducibility of the of the semantic space in SSD, whereas this effect may be diluted in BERT embeddings, which integrate broader contextual information and non-lexical elements. Further research is needed to address this and disentangle the relation the effects of different models and reducibility measures.

Additionally, to gain more insight into the architecture of BERT’s hidden layer model, we estimated ID on a layer-by-layer basis. All the samples and groups showed a similar pattern of increase of ID in the early layers, with a peak at a certain layer, and then declined in the later layers, although differing in the location of the peak. For German and Spanish samples, differences between groups appeared in later layers where semantic relationships are supposed to be encoded, in contrast to the earlier layers that encode more surface-level linguistic features (Jawahar et al., 2019). This also suggest that the reducibility of the embeddings space defined by words in speech occurs at a semantic level, although it may also be affected by grammatical structure.

The notion of semantic reducibility must be distinguished from semantic similarity as previously studied in the literature. While these two phenomena may be related, our correlation analyses revealed that they are not trivially equivalent. For example, the number of components required to explain 90% of variance (*Ncomp*_90_) and *ID* showed strong correlations with each other and with the variance explained by the first two components (*EXVar*_2_), particularly in the clinical groups. However, their correlation with mean semantic similarity was weak and inconsistent across languages. Only in the SSD group of the German sample did *Ncomp*_90_ correlate weakly and negatively with semantic similarity, and only in the English HC group did ID show a weak positive correlation with similarity. These mixed patterns suggest that semantic reducibility captures more than just surface-level similarity between words, and it may reflect the intrinsic geometry of the semantic space of speech, as well as the grammatical constraints that are entangled with specific lexical selections in a particular speech sample. Additionally, to explore further the relationship between semantic similarity and reducibility measures, it would be necessary to compare individuals with same speech lengths and assess whether these associations vary by clinical group.

It is worth considering that the reducibility of the semantic space may partly reflect underlying grammatical constraints. While our analyses focused on content words, lexical selection in speech is tightly coupled with grammar. In this context, the observed geometric properties may not only stem from lexical choices but also from the grammatical scaffolding that shapes them. A possible direction for future research would be to examine the contribution of grammatical features to the reducibility of the semantic space across clinical groups.

Further, clinical status may influence semantic reducibility. For example, in the English sample, the FEP group showed more reducibility compared to HC, while CS patients, who presented relatively mild symptomatology at the time of recording, did not differ significantly from HC. This finding aligns with the idea that reducibility effects are most pronounced during acute or early phases of psychosis, such as FEP, and supports a phase-dependent interpretation, where alterations in the semantic structure of speech are more clearly observed during early illness stages and may diminish with clinical stability or treatment. Lexical reducibility in this context may reflect cognitive rigidity and associative constraint, particularly pronounced in FEP.

Finally, while our study focused on BERT-base and *fastText*, future research should explore whether the same effects hold when using larger and more diverse models, such as LLaMA, Mistral, or Gemma. Expanding this research across more languages and embedding architectures would provide further insights into the cross-linguistic generalizability and computational robustness of the reducibility phenomenon.

## Data Availability

Data used for the present analyses are not at present available publicly, but access within the stipulations laid down by the respective ethics committees can be requested from the local PIs: FS (German), RA (Spanish), LP (English).

